# Māori and Pacific People in New Zealand have higher risk of hospitalisation for COVID-19

**DOI:** 10.1101/2020.12.25.20248427

**Authors:** Nicholas Steyn, Rachelle N. Binny, Kate Hannah, Shaun C. Hendy, Alex James, Audrey Lustig, Kannan Ridings, Michael J. Plank, Andrew Sporle

## Abstract

**Aims:** We aim to quantify differences in clinical outcomes from COVID-19 infection in Aotearoa New Zealand by ethnicity with a focus on risk of hospitalisation.

**Methods:** We used data on age, ethnicity, deprivation index, pre-existing health conditions, and clinical outcomes on 1,829 COVID-19 cases reported in New Zealand. We used a logistic regression model to calculate odds ratios for the risk of hospitalisation by ethnicity. We also consider length of hospital stay and risk of fatality.

**Results:** Māori have 2.50 times greater odds of hospitalisation (95% CI 1.39 – 4.51) than non-Māori, non-Pacific people, after controlling for age and pre-existing conditions. Pacific people have 3 times greater odds (95% CI 1.75 – 5.33).

**Conclusions:** Structural inequities and systemic racism in the healthcare system mean that Māori and Pacific communities face a much greater health burden from COVID-19. Older people and those with pre-existing health conditions are also at greater risk. This should inform future policy decisions including prioritising groups for vaccination.

## Introduction

Up to 25 September 2020, New Zealand reported 1,829 confirmed and probable cases of COVID-19, a disease caused by a novel coronavirus originating in Wuhan, China. The majority of these cases were associated with one of two outbreaks of sustained community transmission: the first in March/April 2020 and the second in August/September 2020. Up to 22 May, there were 1,504 confirmed and probable cases, of which 573 had a recent history of international travel. Between 22 May and 11 August, there were 65 cases, all of which were in detected in international arrivals and contained in government-managed isolation facilities. Between 11 August and 25 September, 260 cases were reported, with the majority linked to a large cluster in Auckland.

The August cluster differed substantially from the initial outbreak in March/April 2020. The vast majority of cases resulted from workplace, community, public transport and household transmission, rather than being associated with international travel ^1^. The August cluster had a higher proportion of cases under 20 years old and a lower proportion of cases over 60 years old than the earlier outbreak (Figure 1). It also contained a much higher proportion of cases among the Pacific and Māori populations than the first outbreak ^1, 2^. Multigenerational living is proportionately greater in Pacific peoples as a population, but the lack of high-quality suitable housing means that this is often overcrowded^3^. Pacific people also experience poorer access to healthcare^4^ and are at greater risk of clinically severe outcomes from COVID-19 infection ^5^.

**Figure 1.**
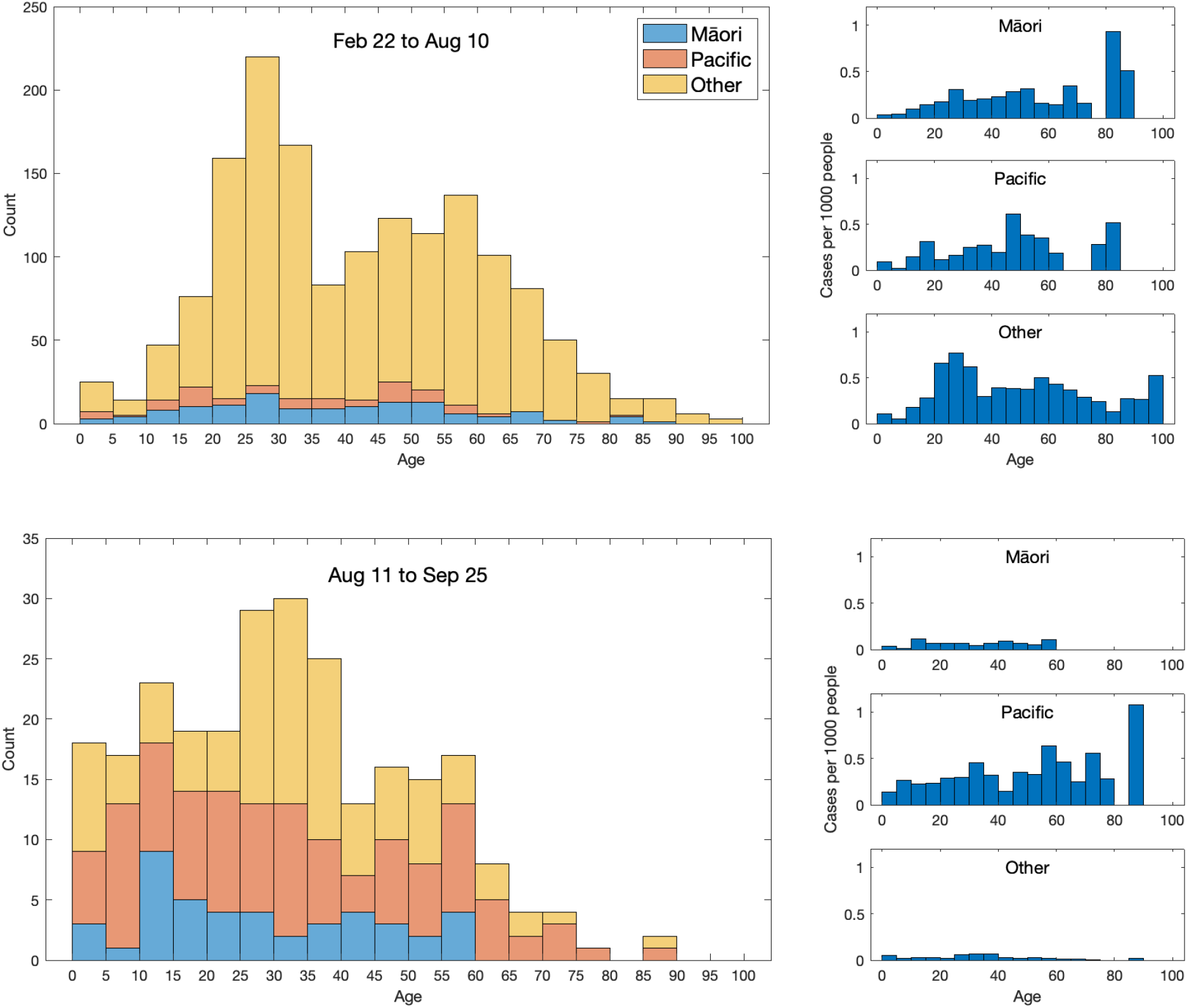
Age-ethnicity structure of New Zealand’s two major outbreaks of COVID-19 using prioritised ethnicity. The plots on the right give the number of cases per 1000 people in that age-ethnicity grouping, population data from Census 2018 ^2^.

Historically, Māori, and Pacific communities both in New Zealand and in the Pacific have had worse experiences of pandemics. During the 2009 H1N1 influenza pandemic, the rate of infection for Māori was twice that of Pākehā, with increased severity ^6^. Our recent research estimated similar inequities would occur in the infection fatality rate for COVID-19 ^5^.

New Zealand’s effective public health response to the pandemic limited the number of COVID -19 fatalities during 2020 to 25 ^7^, which corresponds to a fatality rate of 5 deaths per million people. This means that there is, thankfully, insufficient empirical data at present to reliably estimate differences in the infection fatality rate by ethnicity. Here, we aim to determine whether there are significant differences by ethnicity in the risk of clinically severe outcomes from COVID-19 measured by the hospitalisation rate and length of hospital stay. We take a data-driven approach, using information that is routinely collected for all cases of COVID-19 in New Zealand. The available data are imperfect and the number of cases is relatively small, but it is nonetheless the best data currently available to understand differences in risk from infection with COVID-19 between different ethnicities in New Zealand. The results are important for future policy decisions and pandemic planning, for example identification of priority groups for vaccination against COVID-19.

## Methods

We developed three separate risk models to quantify the risk of hospitalisation, length of hospital stay and fatality risk. Each model used the same methodology and set of predictor variables.

### Data

Case data was obtained from the EpiSurv database on all 1,829 confirmed and probable cases of COVID-19 reported in New Zealand up to 25 September 2020. EpiSurv is New Zealand’s national notifiable disease surveillance database, operated by Environmental Science and Research (ESR) on behalf of the Ministry of Health^8^. EpiSurv collates notifiable disease information on a real-time, including case demographics, clinical features and risk factors. The data for COVID-19 cases includes hospitalisation status and dates, clinical outcome (i.e. recovered, death), age, sex, presence/absence of several underlying health conditions (see below), StatsNZ meshblock of current home address, and self-reported ethnicity (see Table 1). Ethnicity information in EpiSurv is collected on the standard COVID-19 case report form ^8^, where it is described as ‘core surveillance data’ ^9^. The responses are then prioritised to a single response using the Ministry of Health’s Ethnicity Data Protocols ^10^. The ethnicity information in the Ministry of Health sourced data includes multiple ethnicity fields sourced by linking EpiSurv data to the National Health Index (NHI) data collection.

**Table 1.** Summary of case data. Deprivation index was used in its raw index form in the model but has been presented as quintiles for ease of interpretation, with the 1^st^ quintile representing those that reside in a meshblock with the lowest socioeconomic deprivation and the 5^th^ quintile the highest deprivation. Age is also presented in discretised brackets. The use of total ethnicity datameans sums over these rows will be greater than the totals where some cases are recorded as having multiple ethnicities.

|  | Hospitalised | Mean length of stay (days) | Died | Total |
| --- | --- | --- | --- | --- |
| Overall | 120 (6.6%) | 8.2 | 25 (1.4%) | 1829 |
| Ethnicity |  |  |  |  |
| Māori | 18 (10.1%) | 9.4 | 4 (2.2%) | 178 |
| Pacific | 21 (10.0%) | 11.6 | 1 (0.5%) | 210 |
| Asian | 15 (5.0%) | 4.1 | 0 (0.0%) | 300 |
| NZ European/Other | 69 (5.5%) | 7.8 | 21 (1.7%) | 1259 |
| Health Status |  |  |  |  |
| Underlying cond. | 47 (14.2%) | 9.0 | 13 (3.9%) | 330 |
| No underlying cond. | 73 (4.9%) | 7.6 | 12 (0.8%) | 1499 |
| Sex |  |  |  |  |
| Male | 57 (6.9%) | 9.2 | 14 (1.7%) | 823 |
| Female | 63 (6.3%) | 7.3 | 11 (1.1%) | 1006 |
| Age |  |  |  |  |
| 0-19 | 4 (1.7%) | 2.8 | 0 (0.0%) | 239 |
| 20-39 | 23 (3.1%) | 2.6 | 0 (0.0%) | 732 |
| 40-59 | 43 (8.0%) | 8.1 | 2 (0.4%) | 538 |
| 60-79 | 38 (13.6%) | 8.6 | 10 (3.6%) | 279 |
| 80+ | 12 (29.3%) | 19.2 | 13 (31.7%) | 41 |
| Deprivation |  |  |  |  |
| 1<sup>st</sup> quintile (least) | 17 (4.0%) | 10 | 3 (0.7%) | 422 |
| 2<sup>nd</sup> quintile | 35 (8.3%) | 6.6 | 5 (1.2%) | 421 |
| 3<sup>rd</sup> quintile | 11 (3.7%) | 9.2 | 2 (0.7%) | 300 |
| 4<sup>th</sup> quintile | 33 (8.8%) | 6.5 | 1 (0.3%) | 374 |
| 5<sup>th</sup> quintile (most) | 18 (6.5%) | 10.4 | 14 (5.0%) | 278 |
| Missing | 6 (17.6%) | 13.5 | 0 (0.0%) | 34 |

The data on underlying health conditions were simplified into a binary variable indicating if the individual had at least one of the following conditions: chronic lung disease, cardiovascular disease, diabetes, immunodeficiency, asthma, or malignancy. These conditions were chosen because they are all recorded in the EpiSurv dataset^8^ and are known to be associated with increased risk of COVID-19 hospitalisation ^11^. We did not consider the effects of multiple underlying health conditions due to the limitations of analysing such small numbers (see Discussion for limitations associated with this). Of the 1,829 cases, 269 cases (14.7%) had one of the above conditions recorded, 55 cases (3.0%) had two conditions recorded, 4 cases (0.2%) had three conditions recorded and 2 cases (0.1%) had four conditions recorded.

The meshblock number of residential address was used to allocate a measure of geographic socioeconomic deprivation based on the New Zealand deprivation index (NZDep18) ^12^. This was not available for 34 cases, so any models that include deprivation index had a sample size of 1,795 cases and 114 hospitalisations.

Total ethnicity data was used to assign individuals into one or more of the following groups: Māori, Pacific, Asian, NZ European/Other. Due to the limitations of analysing small numbers of cases and to avoid overfitting, we assigned individuals whose ethnicity was recorded as Middle Eastern/ Latin American/African (n=49 cases, 1 hospitalisation) or Other (n=5 cases, 1 hospitalisation) to the NZ European/Other ethnicity group. Individuals for whom total ethnicity data was missing (n=29 cases, 1 hospitalisation) were assigned to the ethnicity recorded in the “prioritised ethnicity” field in EpiSurv. Of the 29 cases with missing total ethnicity data, prioritised ethnicity was recorded as Māori for 1 case, Pacific for 2 cases, Asian for 11 cases and NZ European/other for 15 cases. Of all 1,829 cases, 1,719 (94%) had a single ethnicity recorded, 102 (5.6%) had two ethnicities recorded, and 8 (0.4%) had three ethnicities recorded. A breakdown of the number of cases in the data set by ethnicities is shown in Supplementary Table 1.

Of the 120 hospitalised cases, only 102 had listed discharge dates, which were required for analysis on the length of hospital stay. Five of the 18 cases without discharge dates recorded resulted in death, so their discharge date was set to the date of their death. The remaining 13 cases (two who had not recovered by 25 September 2020 and 11 with no discharge date recorded) were excluded from the length of stay analysis. One additional case was excluded as the discharge date recorded was prior to the hospitalisation date. This resulted in a sample of 106 cases with a recorded length of stay in hospital (see Figure 2). Of the 14 excluded cases, 7 (50%) were in Pacific people, despite Pacific people only making up 18% of hospitalisations. This reduced the sample size for Pacific people and likely biased the results.

**Figure 2.**
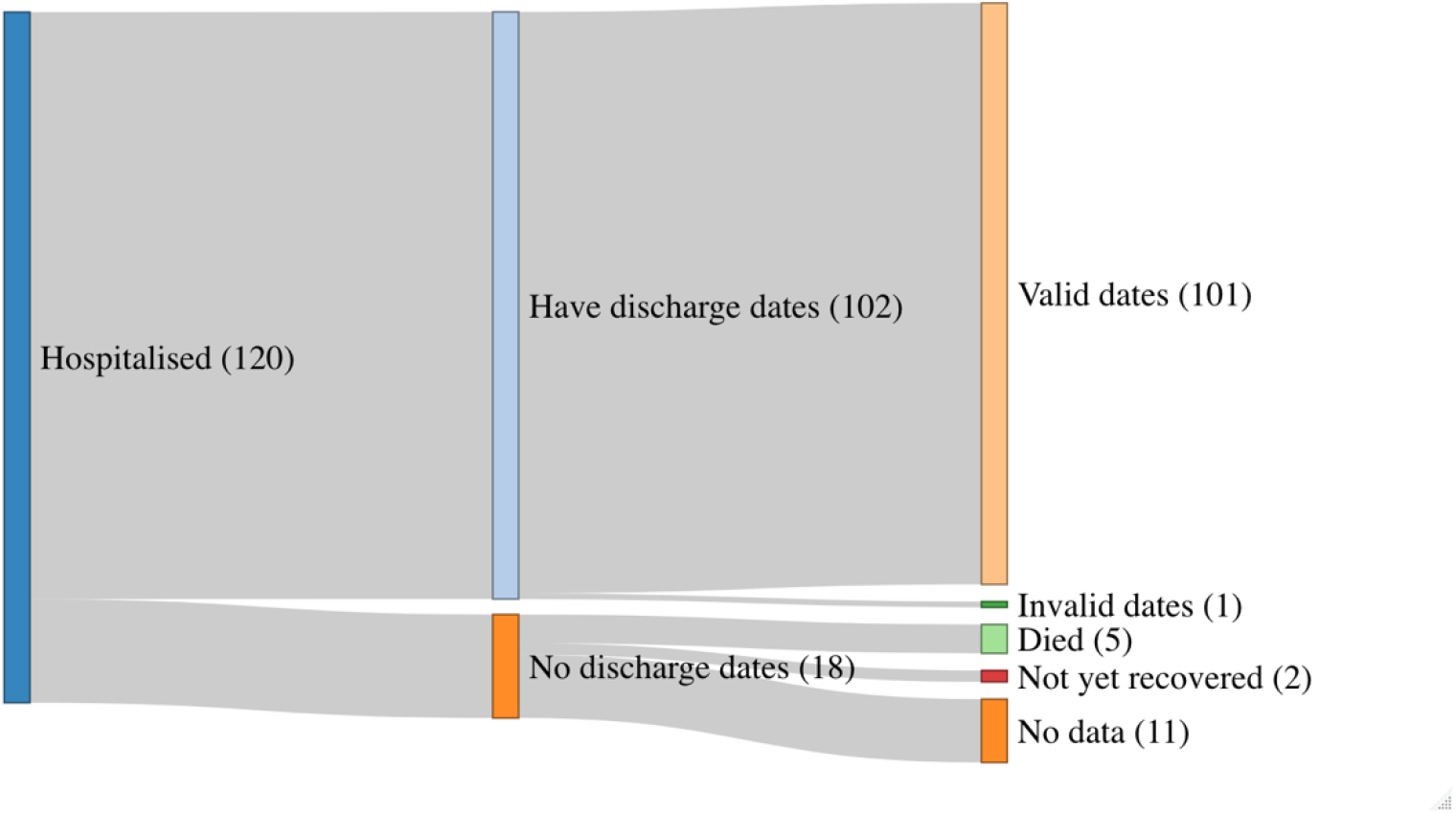
Sankey diagram of case data that was included/excluded from the length of hospital stay analysis. Those with valid discharge dates or death dates (n = 106) were included in the analysis. The remaining cases were excluded (n = 14), of these 1 was Māori, 7 were Pacific, 1 was Asian, 5 were NZ European/other, and 0 had multiple ethnicities recorded. Data are for cases reported up to 25 September 2020.

### Model Selection

For each of the three models, we carried out a simple analysis to determine which predictor variables to include in the model. We used a logistic regression to determine which of ethnicity, underlying health conditions, sex, age, and deprivation should be included. We used Akaike information criterion (AIC) and the area under the receiver operating characteristic curve (AUC) for model selection. Using AIC is a standard, likelihood-based procedure for model selection that quantifies how parsimoniously the model describes the data and penalises models with too many variables ^13^. AUC measures how accurately the model predicts the outcome of interest (in this case hospitalisation) for cases in the dataset ^14^. The complete model was:

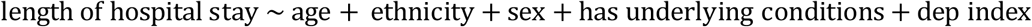

Ethnicity was treated as a categorical variable with individuals belonging to one of Māori, Pacific, Asian, or NZ European/Other. In the case of multiple recorded ethnicities, as there was insufficient data to consider all ethnicity combinations, the standard Ministry of Health prioritisation was used ^10^ for the model selection phase (see below for estimation of effect sizes using multiple ethnicity data). The NZ European/Other group is used as the baseline group so that resulting odds-ratios are interpreted as “difference in risk relative to NZ European/Other”. AIC requires all models to have the same sample size, so during model selection the 34 records with missing deprivation index were removed.

### Estimating the Effect of Ethnicity

During the model selection phase, ethnicity (using prioritised ethnicity) was consistently identified as a significant predictor variable in all three models. Using priority ethnicity neglects important information on individuals who were in multiple ethnicity groups ^15^. For example, there were 19 individuals who were recorded as Māori and Pacific, none of whom were hospitalised. In the standard prioritisation routine, these individuals were classified as Māori and did not, therefore, contribute to model estimates for Pacific people. This undercounted Pacific cases and potentially created age-related biases in the results for Pacific people, as younger Pacific people are more likely to report multiple ethnicities ^16^. To account for this, we reran each model using different ethnicity prioritisation orderings (see Table 2). Odds-ratios and confidence intervals on the odds-ratios were obtained by exponentiating the coefficient estimates and confidence intervals on the coefficient estimates for each risk factor.

**Table 2.** Ethnicity prioritisation ordering depending on the ethnicity effect being estimated.

| Ethnicity effect being estimated | Prioritisation ordering |
| --- | --- |
| Māori | Māori, Pacific, Asian, NZ European/Other |
| Pacific | Pacific, Māori, Asian, NZ European/Other |
| Asian | Asian, Māori, Pacific, NZ European/Other |

### Length of Stay and Risk of Fatality

In addition to the risk of hospitalisation, we used a linear model to consider the effect of these variables on length of hospital stay:

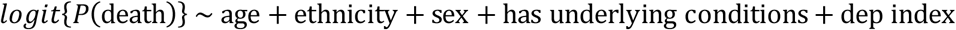

Finally, despite very limited data, we also considered fatality risk under the same framework:

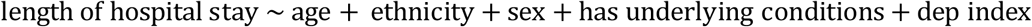

In this final model, as there were no fatalities in Asian people, we combined the Asian and NZ European/Other ethnicity groups. For both these models, we used the same methodology as for the risk of hospitalisation model, i.e. we used standard ethnicity prioritisation to identify significant predictor variables then re-analysed the contribution of these predictor variables under different ethnicity prioritisation orderings.

### Sensitivity analysis

To check how robust our conclusions were to assumptions about ethnicity data and other potential sources of bias, we performed a sensitivity analysis by rerunning the preferred models for risk of hospitalisation and length of hospital stay under each of the following assumptions:

1. Cases with primary ethnicity recorded as Middle Eastern/Latin American/African (n=49 cases, 1 hospitalisation) or Other (n=5 cases, 1 hospitalisation) were excluded from the dataset.
2. Cases with missing total ethnicity data (n=29 cases, 1 hospitalisation) were excluded from the dataset.
3. Cases satisfying either 1 or 2 above were excluded from the dataset.
4. Cases with missing total ethnicity data were assumed to be Māori.
5. Cases with missing total ethnicity data were assumed to be Pacific.
6. Cases with missing total ethnicity data were assumed to be Asian.
7. Cases with missing total ethnicity data were assumed to be NZ European/Other.
8. Cases with a recent overseas travel history (n=707 cases) were excluded from the dataset.
9. Cases with missing length of hospital stay data were assumed to have length of stay 0 days (the shortest stay in the dataset).
10. Cases with missing length of hospital stay data were assumed to have length of stay 52 days (the longest stay in the dataset).

## Results

### Risk of Hospitalisation

For risk of hospitalisation, the model containing age, ethnicity, and the presence of underlying health conditions as predictor variables gave the most parsimonious fit (lowest AIC). This model also has the same predictive power (similar AUC) as more complex models (see Table 3). Including interaction terms did not improve the model fit as measured by AIC. Age was always the strongest predictor of hospitalisation and was included in all models. After age has been accounted for, the best two-variable model also included ethnicity.

**Table 3.** AIC and AUC values for the eight models for risk of hospitalisation with lowest AIC, as well as the age-only and ethnicity-only models. Smaller values of AIC indicate a more parsimonious model fit; larger values of AUC indicate better predictive power.

| Model | AIC | AUC | Model | AIC | AUC |
| --- | --- | --- | --- | --- | --- |
| Age + Eth + HasCond | 755 | 0.762 | Age + Eth + Sex | 761 | 0.758 |
| Age + Eth + HasCond + Sex | 757 | 0.763 | Age + Eth + Dep | 761 | 0.757 |
| Age + Eth + HasCond + Dep | 757 | 0.762 | Age + Eth + Sex + Dep | 763 | 0.758 |
| Age + Eth + HasCond + Sex + Dep | 758 | 0.763 | Age | 779 | 0.728 |
| Age + Eth | 759 | 0.757 | Eth | 844 | 0.577 |

Coefficient estimates associated with sex were always close to zero and had consistently large p-values, indicating that sex was not a strong predictor of hospitalisation in New Zealand’s COVID-19 cases. This is contrary to some international evidence that suggests men suffer worse clinical outcomes on average ^11^. Deprivation index was only statistically significant when considered alongside age but not ethnicity. Deprivation index and ethnicity were slightly correlated, so this suggests the effect of deprivation index was partially captured by ethnicity. Different age groups were represented differently across different levels of deprivation index, suggesting a model containing a deprivation index-age interaction term may be suitable. This was tested and the resulting coefficients were not statistically significant.

Māori and Pacific people are known to have higher rates of multi-morbidity and underdiagnosis of comorbid conditions ^17 18 19^. This suggests that including a term in the model for the interaction between ethnicity and presence of underlying health conditions could be important. However, this term was found to be not statistically significant.

Table 4 and Figure 3 show the results for the preferred model for risk of hospitalisation. Age was associated with a 4.5% increase in odds of hospitalisation per additional year. The presence of at least one underlying health condition increased the odds of hospitalisation by 1.74 times (95% CI 1.14 – 2.65, *p* = 0.01). After controlling for age and underlying conditions, Māori and Pacific people had substantially higher odds of being hospitalised for COVID-19 than other ethnicities: Māori 2.5 times higherodds (95% CI 1.39 – 4.51, *p* = 0.002) and Pacific people 3.06 times higher odds (95% CI 1.75 – 5.33, *p* = 8 × 10^−5^). Asian people were also at higher risk, with 1.35 times higher odds, although this result was not statistically significant (95% CI 0.74 – 2.48, *p =* 0.33).

**Table 4.**
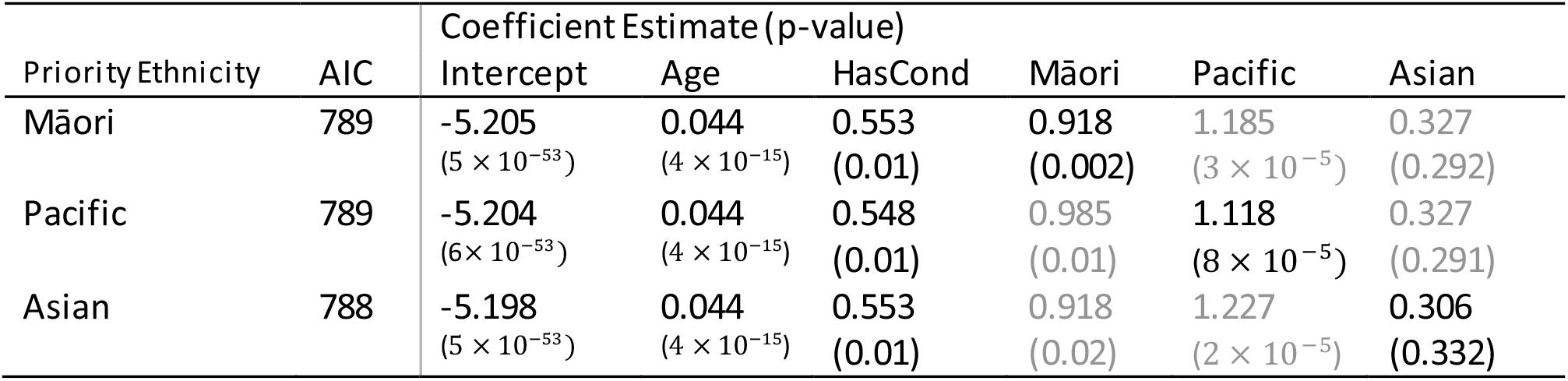
Results of the preferred model (age, ethnicity, underlying conditions) for risk of hospitalisation under each ethnicity prioritisation ordering. Coefficient estimates for ethnicity that is not the priority (grey text) should be treated with caution. These models use the data from all cases (1,829 individuals).

**Figure 3.**
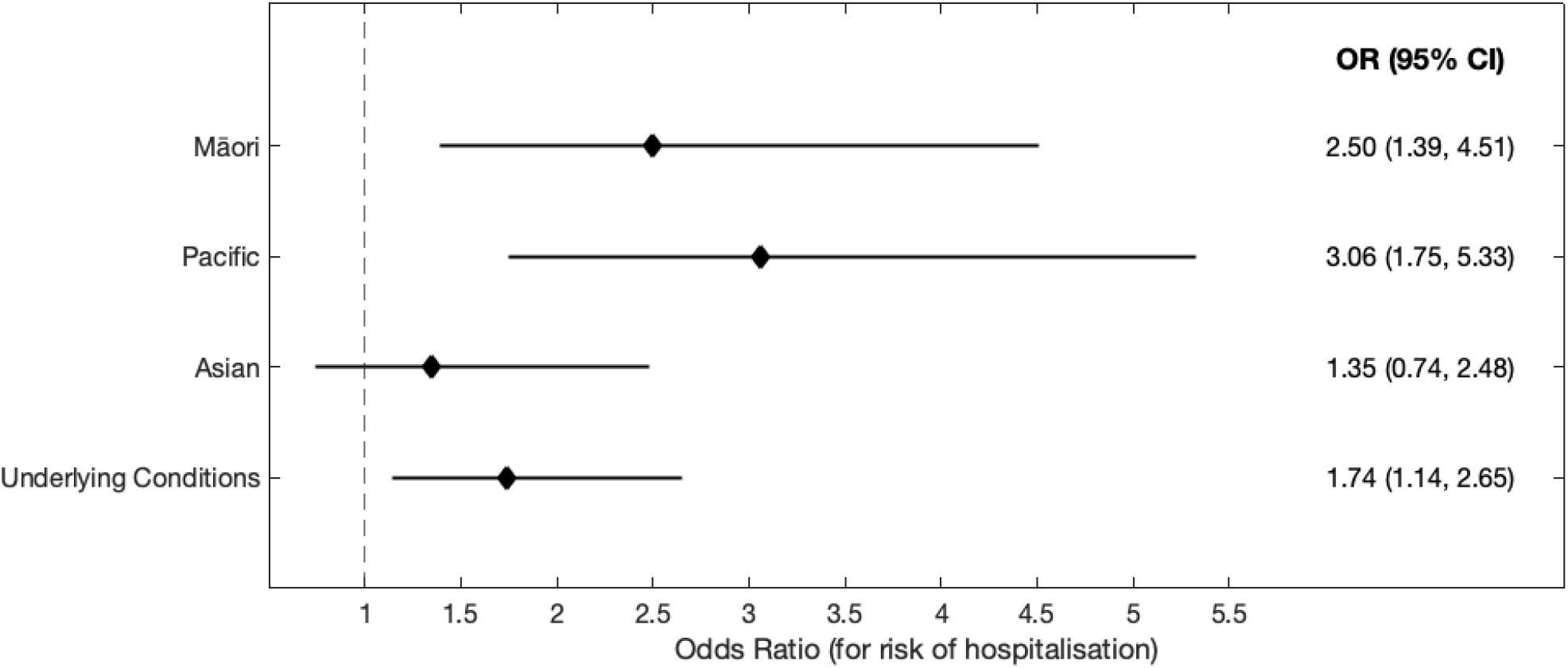
Odds-ratios and 95% confidence intervals for the considered risk factors. The odds-ratio for underlying conditions was taken from the model with Māori as the priority ethnicity, however these results change very little under different prioritisations. In the same model, the odds-ratio for an additional year of age was 1.045 (1.034, 1.057). Analysis based on cases reported up to 25 September 2020.

The odds ratios for different ethnicities shown in Figure 3 represent the increase in risk after controlling for underlying health conditions, which are present in higher rates in Māori and Pacific people ^17^. In the 1,829 cases in the data, there was only a very small correlation between having underlying conditions recorded and either Māori ethnicity (Pearson’s r-squared *r*^2^ = 0.07)or Pacific ethnicity (*r*^2^ = 0.02), so the results were not affected by multi-collinearity in these variables.

The model can be used to estimate the probability of hospitalisation following infection with COVID-19 for an individual of a given age, ethnicity, and presence/absence of underlying health conditions (see Figure 4 and see Supplementary Figure 1 for confidence intervals). It can also be used to estimate the age at which Māori or Pacific cases had the same risk of hospitalisation as those at a specific reference age in the NZ European/Other group, after controlling for the presence or absence of underlying health conditions (Table 5). This shows that, on average, there is a 20.7 year age gap between Māori and NZ European/Other, and a 25.2 year age gap between Pacific and NZ European/Other, at the same level of risk. These estimates should be used with caution as they assume that age has the same proportional effect in each ethnicity (see Discussion for limitations and sources of bias).

**Table 5.** Age differences between ethnicities at the same level of risk of hospitalisation. Each row shows a reference age for NZ European/Other and the corresponding age [95% CI] at which Māori and Pacific people have the same predicted risk of hospitalisation as NZ European/Other. Note that, after controlling for underlying health conditions, the average age difference between NZ European/Other and Māori at the same level of risk is always 20.7 years and the average age difference between NZ European/Other and Pacific people at the same level of risk is always 25.2 years, but the size of the confidence intervals varies slightly with age.

| No underlying health conditions |  |  | At least one underlying health condition |  |  |
| --- | --- | --- | --- | --- | --- |
| NZ Euro/Other age | Māori age with same risk | Pacific age with same risk | NZ Euro/Other age | Māori age with same risk | Pacific age with same risk |
| 60 | 39.3 [26.2, 51.8] | 34.8 [22.7, 45.9] | 60 | 39.3 [24.5, 52.0] | 34.8 [20.3, 47.1] |
| 65 | 44.3 [31.6, 57.0] | 39.8 [28.2, 51.2] | 65 | 44.3 [30.1, 56.9] | 39.8 [26.0, 52.0] |
| 70 | 49.3 [36.8, 62.4] | 44.8 [33.5, 56.5] | 70 | 49.3 [35.7, 62.0] | 44.8 [31.6, 57.1] |
| 75 | 54.3 [42.0, 68.0] | 49.8 [38.6, 62.0] | 75 | 54.3 [41.1, 67.3] | 49.8 [37.1, 62.2] |
| 80 | 59.3 [46.9, 73.7] | 54.8 [43.6, 67.7] | 80 | 59.3 [46.4, 72.6] | 54.8 [42.4, 67.6] |

**Figure 4.**
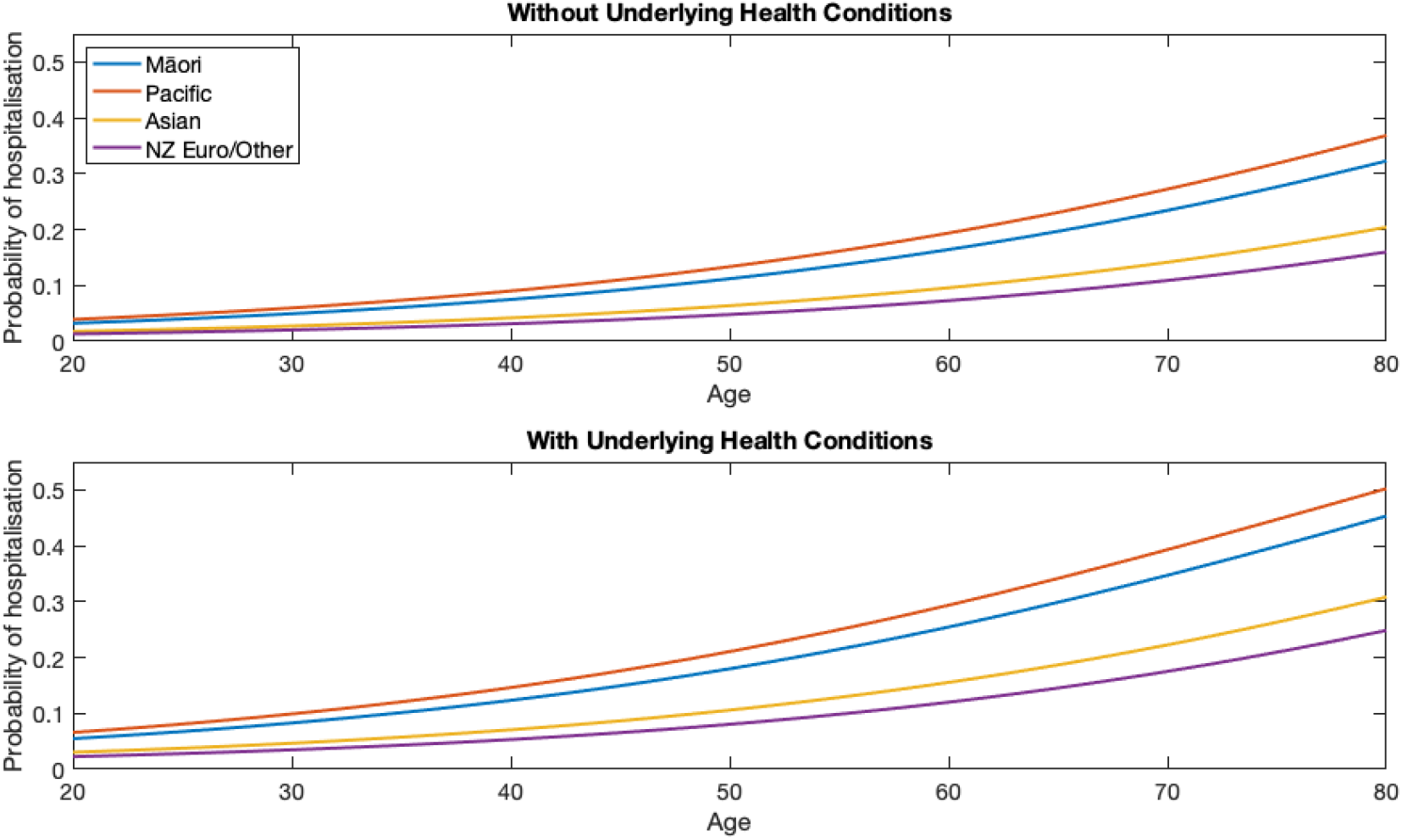
Estimated probability of hospitalisation by age and ethnicity, with and without underlying health conditions. Analysis based on cases reported up to 25 September 2020.

The results of the sensitivity analysis (see Supplementary Table 2) showed that the main conclusions were robust to different assumptions. The magnitude of the odds ratios for Māori and Pacific people could be slightly smaller than those in Figure 3 under different assumptions about missing ethnicity data or ethnicity groupings. For scenarios 1-7 described in Methods, the odds ratio for Māori was always statistically significant and varied between 2.15 (95% CI 1.20 – 3.86) if cases with missing total ethnicity data were assumed to be Māori, and 2.50 (95% CI 1.39 – 4.51) under the default model. The odds ratio for Pacific people was always statistically significant and varied between 2.78 (95 CI 1.61 – 4.80) and 3.06 (95% CI 1.75 – 5.33) under scenarios 1-7. If the EpiSurv ethnicity field (which is more up-to-date but only allows the priority ordering with Māori as priority ethnicity) was used instead of Ministry of Health total ethnicity data, the odds ratio for Māori was 2.68 (95% CI 1.48 – 4.83), which is larger than in Figure 3. Excluding cases with a recent international travel history (scenario 8), increased the odds ratio for Māori and for Pacific people to 2.51 (95% CI 1.28 – 4.93) and 3.20 (95% CI 1.73 – 5.94) respectively. The odds ratio for Asian people was not statistically significant under any of the scenarios tested.

### Length of Hospital Stay

For length of hospital stay, the model containing only age and ethnicity as predictor variables gave the most parsimonious fit (lowest AIC). Age was a more important factor than in the probability of hospitalisation model, with an additional year of age predicting an additional 0.22 days (95% CI 0.14 – 0.31 days, *p* = 2 x 10^-6^) in hospital on average. When used as the priority ethnicity, Māori are expected to spend 4.9 days (95% CI 0.02 – 9.7 days, *p* = 0.052) longer in hospital than NZ European/Other, and Pacific people are expected to spend 5.2 days (95% CI 0.08 – 10.2 days, *p* = 0.049) longer in hospital than NZ European/Other. Length of hospital stay for Asian people was not significantly different from NZ European/Other.

The sensitivity analyses (Supplementary Table 3) showed that the difference in length of hospital stay for Māori was sometimes marginally statistically significant at the *p* = 0.05 level and sometimes not statistically significant, with average length of stay varying between 4.4 days and 6.1 days longer than NZ European. The difference for Pacific people was statistically significant under most scenarios. Under scenarios 1-8 described in Methods, the average length of stay for Pacific people varied between 5.0 days and 5.7 days longer than NZ European. The average length of stay for Pacific people was sensitive to assumptions about cases with missing or invalid length of stay data (scenarios 9-10) because Pacific people were disproportionately represented in this cohort. If cases with missing data were assumed to have length of stay 0 days (the smallest value in the data), the difference in length of stay for Pacific people was not statistically significant. If cases with missing data were assumed to have length of stay 52 days (the largest value in the data), the difference in length of stay for Pacific people was highly significant with an average stay 14.9 days longer than NZ European. These two scenarios are opposite extremes and reality is likely to lie somewhere between them.

### Risk of Fatality

For risk of fatality, the model containing only age and deprivation index as predictor variables gave the most parsimonious fit (lowest AIC). In this model, an additional year of age increased the odds of fatality by 15.9% (95% CI 11.5% – 20.4%, *p* = 3 × 10^−14^). A unit increase in deprivation index was associated with a 0.80% (95% CI 0.33% – 1.27%, *p* = 0.001) increase in the odds of fatality. The difference in deprivation score between the 1^st^ and 4^th^ quintiles in the dataset was 146. This means that the model predicts that an individual at the 80^th^ percentile of deprivation has 3.19 (95% CI 1.62 – 6.31) times the odds of fatality as someone at the 20^th^ percentile in this dataset.

International evidence suggests a linear relationship between log infection fatality rates and age, with one paper estimating an increase in probability of death of 12.9% per year of age ^20^. This is comparable to our results (although changes in the infection fatality rate are not identical to changes in odds, they are close at small probabilities).

The number of fatalities was, fortunately, too small to draw any concrete conclusions on the relationship between risk of fatality and ethnicity. There were no models where ethnicity was a consistently statistically significant predictor of fatality risk. However, this is most likely due to inadequate statistical power of analysing such small numbers. Furthermore, the majority of fatalities are linked to aged care facilities, so are not representative of the type of fatalities if COVID-19 were to become more widespread in the community.

## Discussion

Structural bias and systemic racism are widespread in healthcare systems and are basic determinants of ethnic health inequities in New Zealand and internationally ^4,21^. New Zealand’s experience with the COVID-19 epidemic indicates that Māori and Pacific people are at much greater risk of hospitalisation following infection with COVID-19. It is widely understood from overseas experience that the risk of hospitalisation for COVID-19 increases rapidly with age. However, the effects of ethnicity in New Zealand are not as well understood. Our results show that, an 80 year old case of COVID-19 in the NZ European/Other group without reported comorbidities has the same predicted risk of hospitalisation as a 59.3 year old (95% CI 46.9 – 73.7 year old) case in the Māori group without reported comorbidities. Similarly, an 80 year old case in the NZ European/Other group without reported comorbidities has the same predicted risk of hospitalisation as a 54.7 year old (95% CI 43.6 – 67.7 year old) case in the Pacific group without reported comorbidities. Similar differences are seen across all ages and for cases with at least one reported comorbidity (see Table 6).. These differences in age-specific risk are broadly consistent with earlier estimates of inequities in the COVID -19 infection fatality rate ^5^. Our analysis suggested that average length of hospital stay could be longer for Māori and Pacific people than for NZ European/Other, but the data was insufficient to draw strong conclusions.

**Table 6.** Sources of potential bias and their likely direction of effect on model predictions for the hospitalisation odds ratio (OR) for Māori and Pacific people. **↑** and **↓** indicate that the source of bias is likely to mean that the model underestimates or overestimates respectively the odds ratios for Māori and Pacific people.

| Source of bias and likely direction of effect on hospitalisation OR for Māori and Pacific people |  | Contextual remarks |
| --- | --- | --- |
| Outdated or inaccurate total ethnicity data | ↑ | Using the EpiSurv ethnicity field (which is only prioritised ethnicity using the first priority ordering in Table 2) results a larger estimated OR for Māori. |
| Multimorbidity and underreporting of comorbid conditions | ↓ | Māori and Pacific people have higher rates of multimorbidity and under-reporting of comorbid conditions. If Māori and Pacific cases in the dataset have multiple comorbid conditions or comorbid conditions that are not reported, the reported OR could overestimate the relative risk of hospitalisation. |
| Change in COVID-19 testing over time | ↑ | In the first wave, which was dominated by NZ European cases, testing and contact tracing rates were lower and testing was less accessible than in the second wave, which had more Māori and Pacific cases. If more mild cases were missed in the first wave than in the second, this could make the hospitalisation risk appear lower in the second wave. This could mean the model underestimates the risk for Māori and Pacific people. |
| Change in threshold for hospitalisation over time | — | There is no clear evidence that the threshold for hospitalisation with COVID-19 has changed over time. It is possible that the introduction of hotel quarantine facilities for community cases in the second wave meant mild cases were less likely to be hospitalised. If this were the case, the model could underestimate the risk for Māori and Pacific people. |
| Overrepresentation of international travellers in dataset | ↑ | Excluding overseas cases from the analysis increased the OR for Māori and Pacific people. |

We have only considered the risk of being hospitalised given an individual was infected with COVID-19. The likelihood of hospitalisation will depend on prevailing admission policies in each hospital. These policies may vary across the country and over time, however we ignored this variation in this analysis. The overall risk of being hospitalised also depends on the likelihood of infection, which is specifically not included in our calculations. COVID-19 can spread quickly in communities with higher levels of workplace, community or whānau interaction, crowded housing, insecure employment, and decreased access to healthcare or COVID-19 testing. These are frequently the same individuals, groups or communities that are at higher risk of hospitalisation and fatality if infected, meaning there is additional potential burden of the epidemic on these people.

When fitting each model, we assigned each individual to only one ethnicity as the small number of cases precluded investigation of all combinations of ethnic identity. This means that our results cannot be used multiplicatively to estimate the risk of hospitalisation for an individual belonging to multiple ethnicities. Other effects are multiplicative in the odds. For example, an individual with reported comorbid conditions has odds of hospitalisation that are 74% greater than another individual of the same age and ethnicity without reported comorbid conditions.

We have presented the results of a simple analysis that ignores several potential sources of bias and additional inequities (see Table 6). For example, the recording and the analysis of the effect of comorbid conditions are crude. Different health conditions have significantly different effects and the presence of multiple health conditions may increase risk further. We did not have a sufficient number of cases to estimate the effect of individual health conditions or combinations of conditions. Māori and Pacific people have lower life expectancy, higher rates of multi-morbidity and respiratory illness, higher rates of under-reporting of comorbid conditions, and typically experience adverse health outcomes at an earlier age ^17 18 19^. These factors have not been accounted for in the model and are likely to exacerbate the risk of clinically severe outcomes from COVID-19. It is possible that some of the observed risk of hospitalisation for Māori could be explained by unreported, undiagnosed or multiple comorbid conditions, in which case the odds ratios for ethnicity that we reported could be overestimates.

Testing rates and contact tracing were much higher in the second outbreak in August/September 2020 than in the first outbreak in March/April 2020, meaning that more mild cases of COVID-19 would have been identified in the second outbreak compared with the first. As this second outbreak disproportionately affected Pacific and Māori people, the model may underestimate their relative risk of hospitalisation. Our model is fitted to data from a period in which the prevalence of COVID-19 was low and healthcare services had adequate capacity. Systemic racism within the healthcare system could further exacerbate inequities in outcomes if COVID-19 prevalence increased and healthcare capacity was overstretched ^22 17 23^. Deprivation index was assigned according to the meshblock of an individual’s home address. This may be a good proxy for general current socioeconomic deprivation on average, but the small number of cases in the dataset may not be sufficient for this to apply. Geographic measures of deprivation are widely used and useful as they simply require an address to provide the information. However, such information may not represent the socioeconomic experiences of an individual over their lifetime ^24^.

The level of ethnic group classification used here involved broad categories that define populations with diverse experiences, cultures, nationalities, exposure to racism and immigration histories. The level of ethnicity data available and the absence of migration information(other than recent overseas travel) precluded a more nuanced understanding of the hospitalisation risks within these broad ethnic categories. Understanding the potential impact of the epidemic and informing the delivery of the vaccination programme requires complete and detailed ethnicity information to be included in the routinely available data. This is currently not the case, yet these groups have high risks of poor outcomes from COVID-19 infection. Ideally, ethnicity information should be either collected at the time of testing or sourced from the existing NHI information. The collection of high quality ethnicity information can be done quickly and simply, even in busy clinical settings. This study has also highlighted the differential impact of missing data on understanding the course and impact of the epidemic, which are important for informing interventions including the vaccine delivery programme. Data completeness checks and follow-ups of missing data are simple quality control mechanisms for improving the reliability of routinely collected, but essential information.

The results we have presented are from a relatively small number of cases that may not be representative of the New Zealand population due to the limited spread of these outbreaks. Consequently, although our results are based on all cases for which data is available, caution should be used when generalising the results to other groups or the wider community. The small number of cases and hospitalisations also makes it difficult to separate the effects of different variables, for example the effect of belonging to multiple ethnicities or having multiple comorbid conditions recorded. We have used a likelihood-based approach (AIC) that penalises the use of models with too many variables. The results we have presented are from very simple models that use only two or three predictor variables. This highlights the variables with the largest impacts on the results, but necessarily ignores factors that could have important effects on risk. If in future New Zealand has significantly more hospitalisations from COVID-19, the analysis should be rerun to take account of the additional data. With a larger number of cases, the model selection phase of our approach could include more variables in the model. Our approach uses information that is routinely collected for all cases of COVID-19 in New Zealand, so it would be straightforward to run with an updated dataset.

We conclude that Māori and Pacific people have substantially higher risk of hospitalisation for COVID-19, after controlling for age, presence of underlying health conditions, and socioeconomic deprivation. We have previously estimated that Māori and Pacific people would experience higher infection fatality rates from COVID-19 ^5^. Our new results add to the imperative for New Zealand’s COVID-19 response to include a focus on measures to protect high-risk groups and to prevent the large-scale inequities in health outcomes that would result from widespread community transmission ^25^. Our results also have clear implications for identifying priority groups for vaccination against COVID-19, for which planning is currently underway. They demonstrate that it will be essential to account for ethnicity when targeting vaccination to age groups based on their risk of clinically severe infection.

## Supporting information

Supplementary Tabe 2

## Data Availability

The database of individual cases cannot be made available for privacy reasons.

## Acknowledgements

The authors acknowledge the support of StatsNZ, ESR, and the Ministry of Health in supplying data in support of this work. In particular, we would like to acknowledge Laura Cleary for her work in providing data on total ethnicity and meshblock of home address. The authors are grateful to Melissa McLeod, Ricci Harris, Anja Mizdrak, Patricia Priest, and three anonymous reviewers for comments on an earlier version of this manuscript. This work was funded by the New Zealand Ministry of Business, Innovation and Employment and Te Pūnaha Matatini, Centre of Research Excellence in Complex Systems. Andrew Sporle is also funded by Health Research Council Project Grant 20/1018.

