## Supplementary Text for "Māori and Pacific People in New Zealand have higher risk of hospitalisation for COVID-19"

**Supplementary Material**

**Contents**


- Supplementary Table 1. Ethnicity breakdown of the data.
- Supplementary Table 2 (see supplementary_table2.xlsx). Results of sensitivity analysis for the model for probability of hospitalisation.
- Supplementary Table 3 (see supplementary_table3.xlsx). Results of sensitivity analysis for the model for length of hospital stay.
- Supplementary Figure 1. Model results including 90% confidence intervals for probability of hospitalisation by age and ethnicity.

| | Māori | Pacific | Asian | NZ Euro/other |
|---|---|---|---|---|
| Māori | **99** | 13 | 1 | 59 |
| Pacific | 13 | **166** | 8 | 15 |
| Asian | 1 | 8 | **283** | 6 |
| NZ Euro/other | 59 | 15 | 6 | **1171** |
| Total | 172 | 202 | 298 | 1251 |

**Supplementary Table 1.** Breakdown of the number of cases in the data set by ethnicity showing cases with a single ethnicity (bold numbers) and cases with two ethnicities (non-bold numbers). In addition to the 1,821 single and dual ethnicity cases represented in the table, there were 8 cases with three ethnicities, of which 6 were Māori, Pacific and NZ European/other, and 2 were Pacific, Asian and NZ European/other.

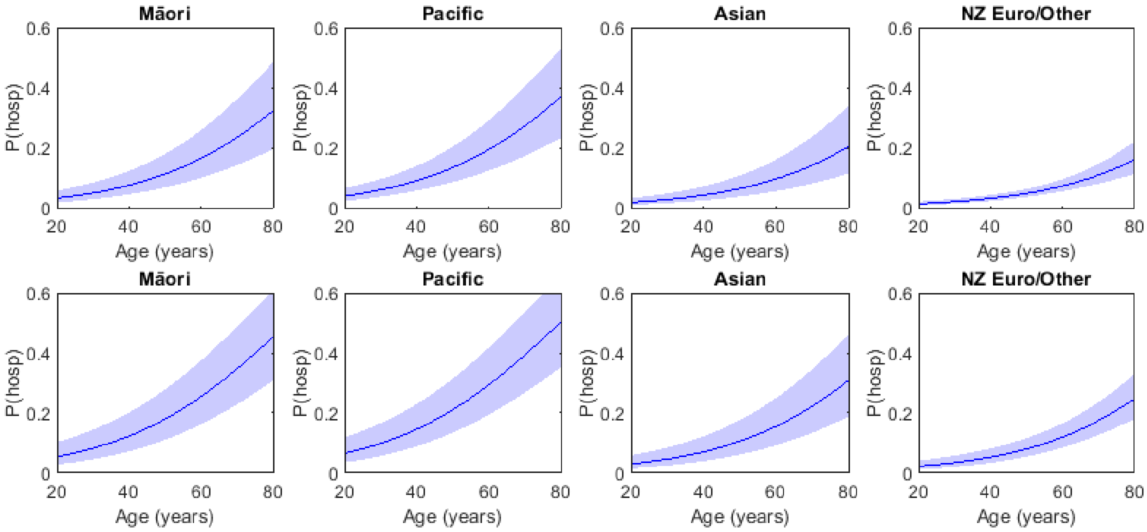


**Supplementary Figure 1.** Model results including 95% confidence intervals for probability of hospitalisation by age and ethnicity for individuals without underlying health conditions (top row of plots) and with underlying health conditions (bottom row of plots).
